# Computational simulation to assess patient safety of uncompensated COVID-19 two-patient ventilator sharing using the Pulse Physiology Engine

**DOI:** 10.1101/2020.05.19.20107201

**Authors:** Jeffrey B. Webb, Aaron Bray, Philip K. Asare, Rachel B. Clipp, Yatin B. Mehta, Sudheer Penupolu, Aalpen A. Patel, S. Mark Poler

## Abstract

**Background:** The COVID-19 pandemic is stretching medical resources internationally, sometimes creating ventilator shortages that complicate clinical and ethical situations. The possibility of needing to ventilate multiple patients with a single ventilator raises patient health and safety concerns in addition to clinical conditions needing treatment.

Wherever ventilators are employed, additional tubing and splitting adaptors may be available. Adjustable flow-compensating resistance for differences in lung compliance on individual limbs may not be readily implementable.

By exploring a number and range of possible contributing factors using computational simulation without risk of patient harm, this paper attempts to define useful bounds for ventilation parameters when compensatory resistance in limbs of a shared breathing circuit is not possible. This desperate approach to shared ventilation support would be a last resort when alternatives have been exhausted.

**Methods:** A whole-body computational physiology model (using lumped parameters) was used to simulate each patient being ventilated. The primary model of a single patient with a dedicated ventilator was augmented to model two patients sharing a single ventilator. In addition to lung mechanics or estimation of CO_2_ and pH expected for set ventilation parameters (considerations of lung physiology alone), full physiological simulation provides estimates of additional values for oxyhemoglobin saturation, arterial oxygen tension, and other patient parameters. A range of ventilator settings and patient characteristics were simulated for paired patients.

**Findings:** To be useful for clinicians, attention has been directed to clinically available parameters. These simulations show patient outcome during multi-patient ventilation is most closely correlated to lung compliance, oxygenation index, oxygen saturation index, and end-tidal carbon dioxide of individual patients. The simulated patient outcome metrics were satisfactory when the lung compliance difference between two patients was less than 12 mL/cmH_2_O, and the oxygen saturation index difference was less than 2 mmHg.

**Interpretation:** In resource-limited regions of the world, the COVID-19 pandemic will result in equipment shortages. While single-patient ventilation is preferable, if that option is unavailable and ventilator sharing using limbs without flow resistance compensation is the only available alternative, these simulations provide a conceptual framework and guidelines for clinical patient selection.

## Introduction

Even in well-resourced healthcare systems the current COVID-19 (SARS-CoV-2 disease) pandemic has led to equipment shortages in Europe, the United States, and is a more severe challenge across the world. Inadequate availability of ventilators for patients in respiratory failure is especially challenging [1-3].

Ventilator sharing or splitting is one mitigation strategy that has been used in practice to rescue as many patients as possible when sufficient numbers of ventilators are not immediately available. Media reports from Las Vegas, Nevada, USA in October 2017 popularized this tactic employed during the surge of over 500 casualties after a mass shooting incident. Manual and improvised ventilation support was needed until stockpiled surge reserve ventilators could be delivered. Curiously, though news media reports are still readily discoverable, no citable healthcare literature directly addressing reported shared ventilator use after the Las Vegas event has been found using extensive searches of PubMed, Google Scholar, and OVID.

The simplest approach for multiplex ventilation is to connect branched tubing without any compensating resistance in any branch for limiting flow to the most compliant patient(s). This is the only way shared ventilation is likely to be implemented rapidly with available tubing and adapters, without ready availability of flow compensating devices.

Only limited testing and clinical experience are available to expose the potential pitfalls of simplistic implementation. Branched breathing circuits to ventilate more than one patient with a single ventilator or compressed gas sources were published by Sommer et al. in 1994 [4], after contemplation of needs for ventilation of mass casualties from gas warfare at the time of the 1990-1 Gulf War [5]. A simpler concept published in 2006 with rudimentary simulation was employed at Las Vegas [6,7]. Tonetti et al. tested feasibility using test lungs of this simply assembled circuit without resistance compensation and tabulated procedures to guide use [8]. Chatburn et al. have studied ventilation of two lung simulators using pressure control and volume control modes of ventilation for six pairs of simulated patients without compensating flow resistance. Simulated carbon dioxide tensions were estimated and used to estimate pH for each modeled patient [9]. Hermann et al. have studied pressure and volume ventilation modes using a computational model of lung ventilation and by test lung experimental simulation [10].

Paladino et al. published a 12-hour proof-of-concept using a single ventilator for four healthy adult sheep, reporting some difficulties even with well-matched “patients” [11]. Letters in response to Paladino’s study raised concerns that the demonstration was not a sound foundation for translation to clinical use, especially for anticipated mass casualty scenarios producing a variety of lung pathophysiologies. They discussed anticipated issues including differing compliances, spontaneous breathing, dyssynchrony with the ventilator, expected shortages of pharmaceuticals and oxygen, monitors, and skilled personnel for vigilant management of patients on shared breathing circuits in a surge situation [5,12].

Due to the perceived risks of having multiple patients per ventilator a consortium of North American professional societies issued a joint statement opposed to this practice, available from several internet sources [13]. The joint statement gives only slight acknowledgment to desperate triage situations whereby dedication of one ventilator exclusively to one patient may determine the demise of another if some means of ventilation cannot be provided. The enumerated considerations of the joint consensus statement are realistic and concerning. However, they abdicate the possibility that split ventilation could be safely and effectively implemented. The identified issues may be considered an agenda for further investigation.

Faced with mounting needs for ventilator support of COVID-19 patients and limited resources, the Federal Emergency Management Administration (FEMA) and healthcare systems prepared clinical guidelines for ventilator sharing using pressure support mode, pharmacological paralysis, and vaguely stated “similar mechanical support needs” [14-16]. Potential risks are enumerated, including unintentional extubation of one patient, cross-contamination, and delayed observation of hypo- or hyper-ventilation. The US Department of Health and Human Services (HHS) issued a document containing a statement that the FDA would not object to split ventilation for the duration of the declared COVID-19 emergency when demand for invasive ventilation exceeds the supply of ventilators [17]. Protocols from a Federal Emergency Management Administration (FEMA) Washington DC COVID-19 Taskforce (Babcock et al. [14]) and the Columbia University College of Physicians & Surgeons collaboration with New York-Presbyterian Hospital (Beitler et al. [15]) are included within the attached HHS document.

Limited clinical demonstrations have been reported. Simple branched-flow circuits for a ventilator do not incorporate flow compensation to limit excessive distribution of ventilation to the most compliant lungs. Beitler et al. reported encouraging results for 3 pairs of carefully matched stable New York COVID-19 ARDS patients, each pair slowly and cautiously prepared for tandem ventilation under a detailed protocol. Each pair was ventilated for 2 days using different equipment or refinements to protocol. Limitations of an anesthesia machine as a ventilator for two patients and consequences of inadequate neuromuscular blockade are reported [18]. Another New York group at Mount Sinai empirically compensated for the maldistribution of ventilation by incorporating a needle valve to restrict breath flowing to the most compliant lungs. Initial simulation testing of shared ventilation using a single ventilator employed two high-fidelity lab manikins. Subsequently, two consented ICU patients were supported for one hour by a single ventilator with a split breathing circuit. At another time, two other consented ICU patients were supported for an hour by one ventilator. Some of the assertions of the previously cited joint statement are addressed by Levin et al. in an appendix [19]. Similar apparatus has been described by an on-line collaborative [20] and another pre-publication document from Yale [21]. Clarke et al. have reported using simple compression clamps on segments of tubing to increase resistance [22].

Han et al. have empirically demonstrated advantages of a somewhat more complicated bag-in-box design for parallel co-ventilation of test lungs driven by a single ICU ventilator, avoiding respired gas cross-contamination, and allowing somewhat independent control of VT, PEEP, and FiO_2_ for widely different lung compliances and single circuit faults [23].

The joint statement and limited empirical information suggest a range of inadequately addressed issues. These provide opportunities for future conventional research, modeling and simulation. However, the pace of conventional research is too slow when addressing a pandemic crisis. Characterizing “last resort” split-circuit ventilation raises complex considerations and questions needing urgent answers to provide “guard rails” parameters for reasonably safe use.

Computational simulation provides an opportunity to quickly develop guidance over a wide range of possible clinical scenarios, without incurring patient risk. While empirical reports have been published [19], we have found only limited reports of empirical human simulation (using masks on volunteers and hyperventilation to avoid spontaneous breathing dyssynchrony) [24], and no computational simulation literature to inform multiple patient shared ventilation, nor specific definition of “similar mechanical support needs” [17]. Thus, it is clear that a method is needed to study and navigate this multi-parameter space and multiple scenarios in this complex system. We undertook a computational simulation of dual patient ventilation with a single ventilator using a computational whole-body simulation model to produce informed clinical parameters addressing noted assumptions and assertions. Use of whole-body physiological simulation exposes secondary effects that would not be apparent when modeling mechanics of ventilation alone.

The Pulse Physiology Engine (Kitware, Inc., Clifton Park, NY) open source software was previously designed and validated to model a single patient-ventilator pairing [25]. We modified and employed this model to simulate two or more patients sharing a single ventilator resource. This computational model permits rapid evaluation of a wide range of possible scenarios to educate clinicians about the potential risks for mismatched patients ventilated through uncompensated branched breathing circuits attached to a common ventilator. As a computational model without risk to patients, clinically untenable possibilities can be explored to determine guidance or “guard rails” to inform clinical practice, hopefully averting risky clinical practices and potentially poor outcomes.

## Methods

### Pulse overview

Concurrent pressure controlled ventilation of two patients in a configuration without flow-compensating resistance is being modeled by computational simulation. The Pulse Physiology Engine (pulse.kitware.com) models are built on the pioneering work of Ty Smith [26], further developed by the Department of Defense as BioGears (W81XWH-13-2-0068), and finally forked, continually developed, and supported for diverging uses as Pulse. Pulse is comprised of lumped-parameter models, which use electrical circuit analogues (e.g., resistors and capacitors) to represent the behavior of physiological regions or systems of the human body [25]. Feedback mechanisms and interactions of systems are accomplished through circuit connections or scalable circuit elements [27]. These circuit models are then solved using transient circuit analysis. Substances can be circulated through the models using pharmacokinetic (PK) and pharmacodynamic (PD) models, which use the physiologic properties of the patient and the physicochemical properties of the drug in differential equations to represent drug diffusion and distribution in the body [28]. Disease and treatment models are designed with differential equations describing the effects of disease and treatment and then applied to the lumped-parameter models affecting the overall calculations. Thus, the Pulse Physiology Engine models patient physiology interacting with medical equipment. The Pulse engine has a rigorous validation standard driven by data from the literature, patient data, and our relationships with clinical experts in a number of medical fields. More details on the transient circuit analysis, substance transport, and validation process can be found in the supplemental data published on the Pulse website (https://pulse.kitware.com/_docs.html). For this study, the Pulse ventilator implementation has been improved. The Pulse lung models were also modified and extended to enable simultaneous simulation of more than one patient interacting with a single ventilator.

### Patient model

Recent studies of laboratory-confirmed COVID-19 pneumonia patients that received invasive mechanical ventilation have shown the PaO_2_/FiO_2_ ratios were consistent with the Berlin criteria for moderate-to-severe Acute Respiratory Distress Syndrome (ARDS) [29,30]. However, in the earliest reports some COVID-19 patients seemed to present an atypical form of ARDS with dissociation between well-preserved lung mechanics (i.e., compliance) and unusually severe pulmonary shunt fraction with consequent hypoxemia [31,32]. Subsequently others have reported typical ARDS respiratory mechanics and gas diffusion parameters for COVID-19 ARDS patients [33,34].

Based on these data, the existing Pulse ARDS pathophysiology model was used. Additionally, the patient airway resistance, compliance, alveolar surface area, and pulmonary shunt fraction can be modified individually or by specifying a lumped-parameter patient disease “severity.” The severity is defined between zero (no change from the healthy state), and one (life-threatening). Severity values of 0.3, 0.6, and 0.9 correspond to accepted criteria for mild, moderate, and severe ARDS, respectively, including PaO_2_/FiO_2_ and shunt fraction changes. To model COVID-19, the ARDS disease severity mappings that represent the combined effects of the reduction in alveolar surface area and increase in pulmonary shunt were used. The combined changes are defined here as the diffusion impairment factor (DIF). The DIF selectively hinders gas exchange and emulates hypoxemic respiratory failure.

Using Pulse’s whole-body modeling approach, the patient is pharmacologically paralyzed simulating use of a neuromuscular transmission blocking agent and endotracheally intubated, consistent with current clinical guidelines [17]. The patient’s computed diseased pulmonary steady state is achieved while also evaluating other effects on the Pulse whole-body model. Computational steps simulate every 2 milliseconds. If a steady state cannot be achieved, those model parameters are rejected.

### Multi-patient ventilator model

The Pulse ventilator model was first developed and tested with single-patient simulations and has been validated through pure simulation and hybrid testing with physical systems [35]. For this study, the Pulse ventilator model was executed in Pressure Control Continuous Mandatory Ventilation (PC-CMV) mode, following existing multi-patient ventilation guidelines [17,21]. A square driving pressure waveform was specified by user-defined Positive End Expiratory Pressure (PEEP), Peak Inspiratory Pressure (PIP), respiratory rate (RR), and Inspiratory: Expiratory (IE) ratio. The ventilator gas fractions, such as Fractional inspired oxygen (FiO_2_), were also specified. The inspiratory, expiratory, and endotracheal tubes are included as part of the lumped-parameter circuit in Pulse. These comprise a closed-loop circuit and a branch for each patient and the ventilator for pneumatic and substance transport analysis. All tubing was estimated to be 3 feet long with a 22 mm inner diameter. The associated tubing resistance is negligible compared to the patient’s tracheobronchial resistance. One-way (check) valves are present to prevent backflow.

Previously, the Pulse engine represented a single patient and the associated equipment. To simulate a single ventilator connected simultaneously to two or more Pulse patients, required the creation of a new multi-patient ventilation engine. The new Pulse multi-patient engine simulates multiple patient physiology engines in lockstep to compute the effects of unequal, and potentially dynamic, differences in patient breathing mechanics when connected in parallel branched breathing circuits subject to a single pressure-mode ventilator. Fig 1 shows an overview of the multi-patient simulation environment.

**Fig 1.**
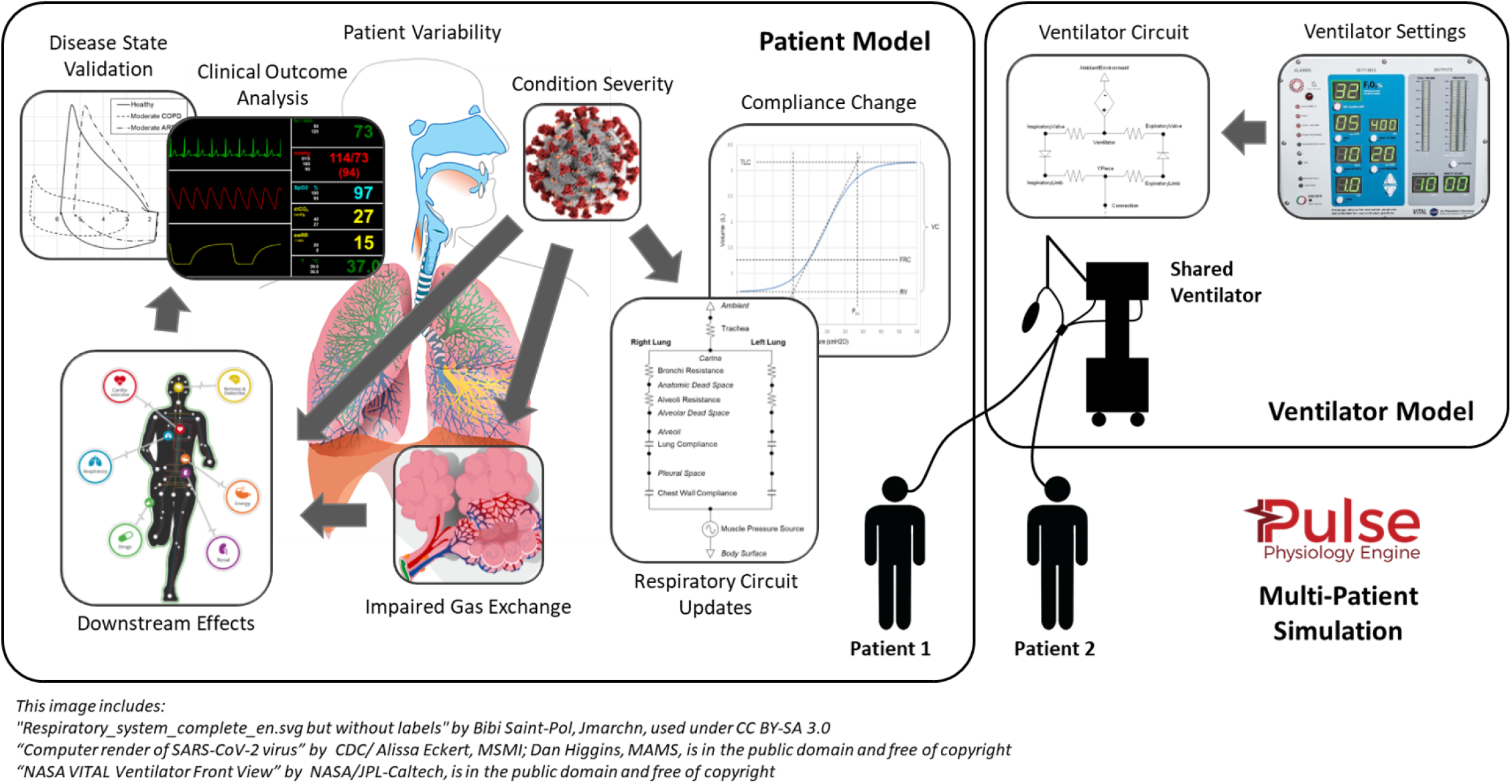
The approach for simulating multi-patient ventilation. An entire system state is calculated every 2 ms. The Pulse dynamic circuit solver and transporter are leveraged to ensure sound physics-based results with conservation of energy and mass. Mechanistic interactions occur with all other Pulse physiological systems, most notably, the alveolar-capillary partial pressure gradient diffusion gas exchange with the cardiovascular system.

### Multi-patient ventilation simulation design

COVID-19 respiratory mechanics parameters were gleaned from the available literature. An initial in silico investigation (computational simulation) used a sparse sampling of the combined mechanical ventilator and COVID-19 patient parameter space to understand patient outcome patterns. These simulations combined various ventilator PEEP settings with patient compliance and disease severity values. Tidal Volume (TV) and arterial partial pressure of oxygen (PaO_2_) were evaluated. This simulation was implemented as a two-step process: 1) patients were simulated with various levels of disease states connected to an individually dedicated ventilator with increasing FiO_2_ values until a homeostatic point (pulse-oximetric oxygen saturation, SpO_2_ > 89%) was reached; then 2) simulated patients were paired for three separate simulations using each patient’s individual ventilator settings and average values for the two paired patients. In the initial model hyperoxia was considered an undesirable, exclusionary outcome, judged to be acceptable for subsequent simulation criteria.A scoring methodology was developed to analyze the simulation results and to identify safe clinical “guard rails” for patient pairing and specifying ventilator settings. The patient pair outcome was scored as follows:

- Positive (green)
  – TV (mL/kg) - both patients between 5.5 and 6.5
  – SpO_2_ (%) - both patients above 89
  – PaO_2_ (mmHg) - both patients below 150
- Less Positive (yellow)
  – TV (mL/kg) - both patients between 4.5 and 7.5
  – SpO_2_ (%) - both patients above 89
  – PaO_2_ (mmHg) - both patients between 150 and 200 (hyperoxemia)
- Negative (red)
  – TV (mL/kg) - either patient below 4.5 (underinflation) or above 7.5 (overinflation)
  – SpO_2_ (%) - either patient under 89 (severe hypoxia)
  – PaO_2_ (mmHg) - either patient above 200 (oxygen toxicity)

If all parameters are in the target range, the result is scored green. If a patient has a tidal volume or SpO_2_ parameter in the yellow or red criteria, the patient is scored in the lowest scoring category. Only in the initial analysis was hyperoxia scored yellow or red.

Results using the first model specification informed a refined discretization of the parameter space in the final simulation study. The initial experiments revealed that the PIP and FiO_2_ should be specified as values providing the best chance of positive outcomes for both patients, as opposed to using average values or those of a single patient. Therefore, the ideal drive pressure (PIP - PEEP) for each pair of patients was directly calculated based on a target tidal volume (6 mL/kg) and known average static respiratory compliance (C_stat_). The FiO_2_ was determined through simulation by finding the lowest value between 0.21 and 1.0 that stabilized both patients to a SpO_2_ of at least 89%. In this final analysis, hyperoxia (oxygen toxicity risk) alone was not scored yellow or red, being considered a likely acceptable expression of good or improving gas exchange.

All possible combinations of patient parameters for the final model required evaluation of 12,642 unique patient and ventilator combinations defined by the following parameter space:

- Mechanical ventilator (one per simulation):
  – Respiratory rate (bpm) - Fixed at 20
  – I:E ratio - Fixed at 1:2
  – PEEP (cmH_2_O) - range of 10 to 20 in increments of 5
  – PIP (cmH_2_O) - Derived from 6 mL/kg given PEEP
  – FiO_2_ - Derived through simulation for both patients’ SpO_2_ > 90% (if possible)
- Patient (two per simulation):
  – Total respiratory resistance (cmH_2_O-s/L) - Fixed at 5
  – Total respiratory static compliance (mL/cmH_2_O) - a range of 10 to 50 in increments of 1
  – DIF (severity 0 to 1) - 0.3 to 0.9 in increment of 0.1

Two equal-sized 70 kg patients are modeled. This should not limit the applicability of these results because pressure mode ventilation does not enforce delivery of a set volume, which could be injurious. Only a single respiratory resistance per patient branch was modeled because clinical variations of airway resistance have not been found to be a prominent characteristic of COVID-19 ARDS [18,29,31,33,34].

## Results

### Patient simulation

This Pulse COVID-19 model is an extension of the existing ARDS methodology that has been validated with referenced empirical data and trends [36]. Scenarios with the Pulse standard, properly ventilated ARDS patient shows good agreement with expected outcomes [37,38]: severities matching mild, moderate, and severe cases resulting in PaO_2_/FiO_2_ of 340, 130, and 50 mmHg, respectively, and shunt fractions of 8%, 27%, and 59%, respectively. Validated ARDS and mechanical ventilator models, with the added ability to specify the patient’s respiratory compliance, allows for reasonable COVID-19 pathophysiology simulations.

Each of the 287 unique COVID-19 patients created (based on DIF and compliance combinations) for the full study were simulated as single patients with their own ventilators to ensure they were capable of achieving 89% SpO_2_ (green outcomes) at maximum FiO_2_. Those patients that were unable to meet this criterion were excluded from further multi-patient ventilation analysis to prevent skewing resulting guidelines. S1 Table shows that 71% of patients with 0.8 DIF and 100% of patients with 0.9 DIF were disqualified for shared ventilator simulation.

### Multi-patient ventilation simulation

Results of an example paired patient simulation are shown in Fig 2. The TV is significantly different because of the C_stat_ mismatch (plot a). The PaO_2_ values and response to oxygenation differ largely from the DIF (plot b). Because CO_2_ transits tissue much more readily than oxygen, the DIF affects O_2_ more severely than CO_2_ (plot b).Time {s)

**Fig 2.**
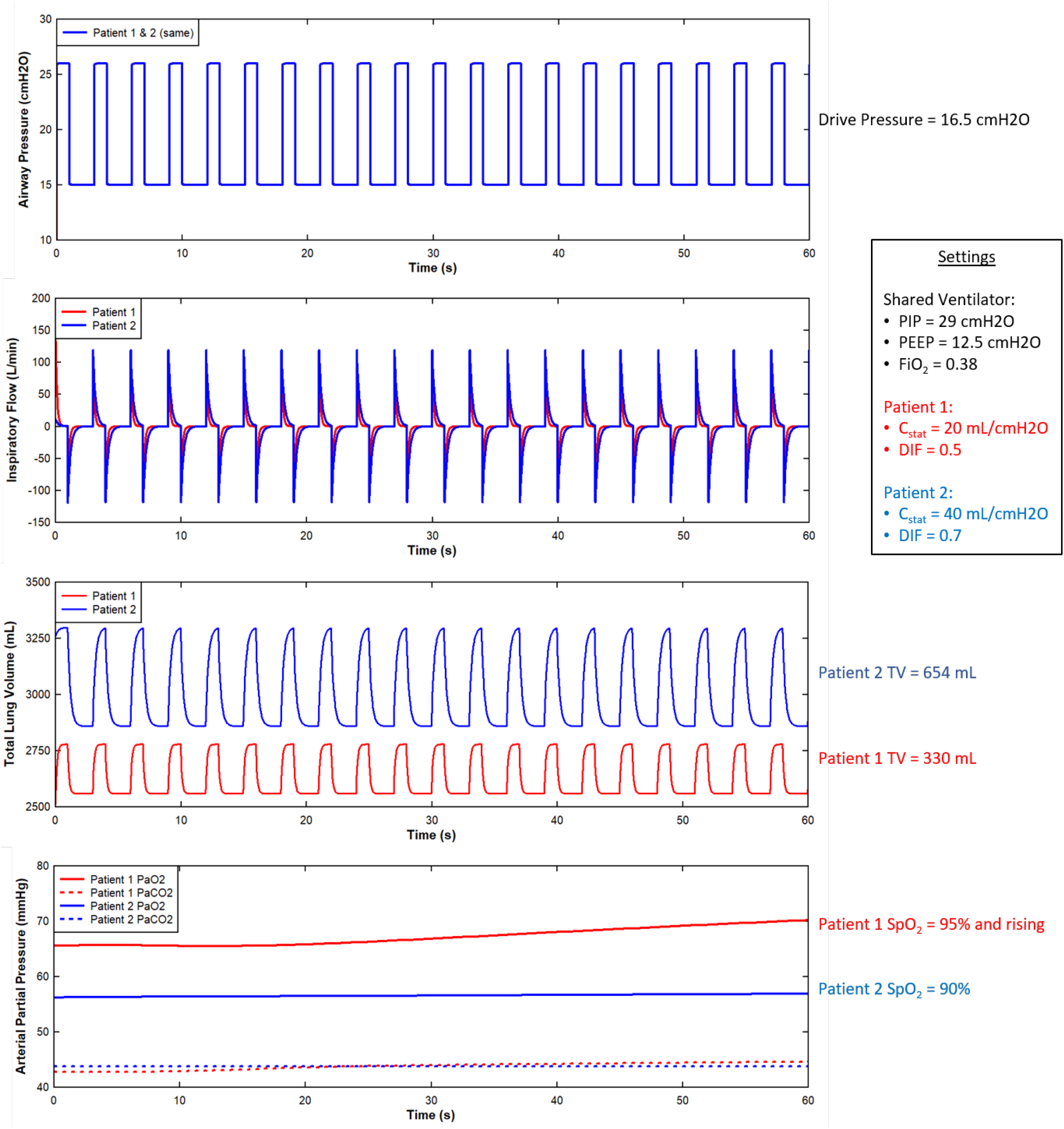
Select outputs from an example multi-patient ventilation scenario from the initial investigation. The outcome of this scenario was classified as negative (red) because both tidal volumes (as mL/kg) are outside the desired bounds for lung protective ventilation.

Parameter differences between patient pairs were evaluated using the scoring methodology described previously. The absolute value of the difference of specific parameters from patient one and patient two was analyzed using a one-way analysis of variance (ANOVA) to determine relative correlations to outcomes. An F-test was used to statistically test the equality of means, with larger *η*^2^ values denoting higher correlation, as shown in Table 1. Five parameters were found to be most correlated to outcomes: C_stat_, End-tidal CO_2_ (EtCO_2_), Alveolar-arterial (A-a) gradient, oxygen saturation index (OSI), and oxygen index (OI). Three of these parameters can be evaluated non-invasively for patients: C_stat_, EtCO_2_, and OSI.

**Table 1.** The results of an outcome correlation statistical analysis. Parameters are differences (Δ) between co-ventilated patients. Larger *η*^2^ values denote greater correlation.

| Parameter | Abbreviation / Equation | Outcome Correlation ($\eta^2$) |
| --- | --- | --- |
| Respiratory Compliance* | $\Delta C_{stat}$ | 0.46 |
| End Tidal Carbon Dioxide* | $\Delta EtCO_2$ | 0.46 |
| Alveolar-arterial Gradient | $\Delta A-a \text{ Gradient} = \Delta(PAO_2 - PaO_2)$ | 0.18 |
| Oxygen Saturation Index* | $\Delta OSI = \Delta(FiO_2 * MAP * 100 / SpO_2)$ | 0.11 |
| Oxygen Index | $\Delta OI = \Delta(FiO_2 * MAP * 100 / PaO_2)$ | 0.11 |
| P/F Ratio | $\Delta P/F \text{ Ratio} = \Delta(PaO_2 / FiO_2)$ | 0.08 |
| Diffusion Impairment Factor<sup>+</sup> | $\Delta DIF$ | 0.02 |
| Mean Airway Pressure* | $\Delta MAP$ | 0.02 |
| S/F Ratio* | $\Delta S/F \text{ Ratio} = \Delta(SpO_2 / FiO_2)$ | 0.02 |

\*Denotes non-invasive, <sup>+</sup>denotes a model parameter

Parameter independence was assessed using the Pearson product-moment correlation coefficient (PPMCC) method. The results in Fig 3 show that two of the four correlated parameters are both non-invasive and independent, C_stat_, and OSI. Therefore, these two parameters were plotted against simulated clinical outcomes in Fig 4.

**Fig 3.**
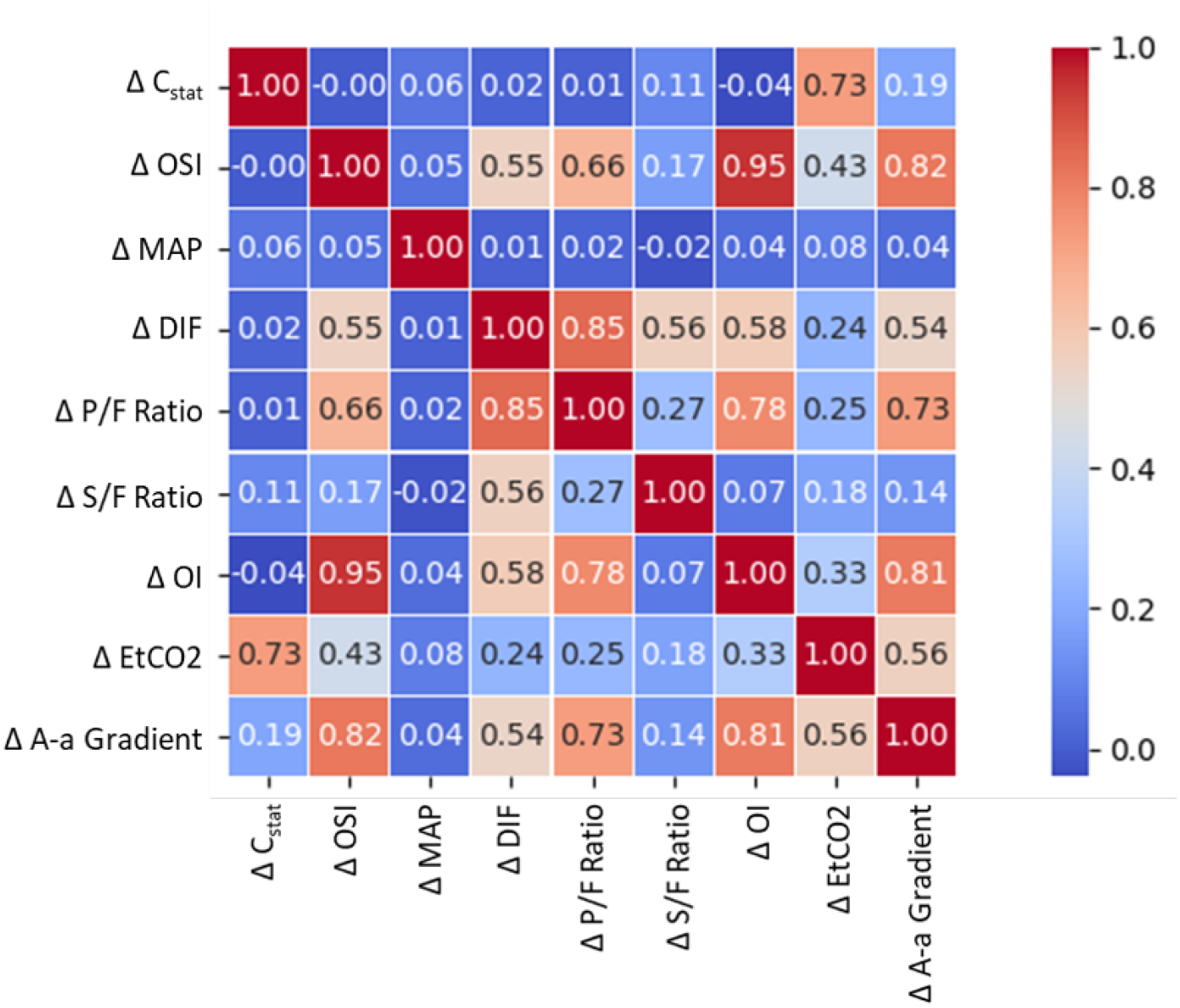
The selected parameters for investigation are compared with each other to determine their dependence. The PPMCC method is used to calculate a value between −1 (inversely correlated) and 1 (correlated). Those with low correction (close to 0) are more independent of each other and are therefore the best candidates for informed decision-making.

**Fig 4.**
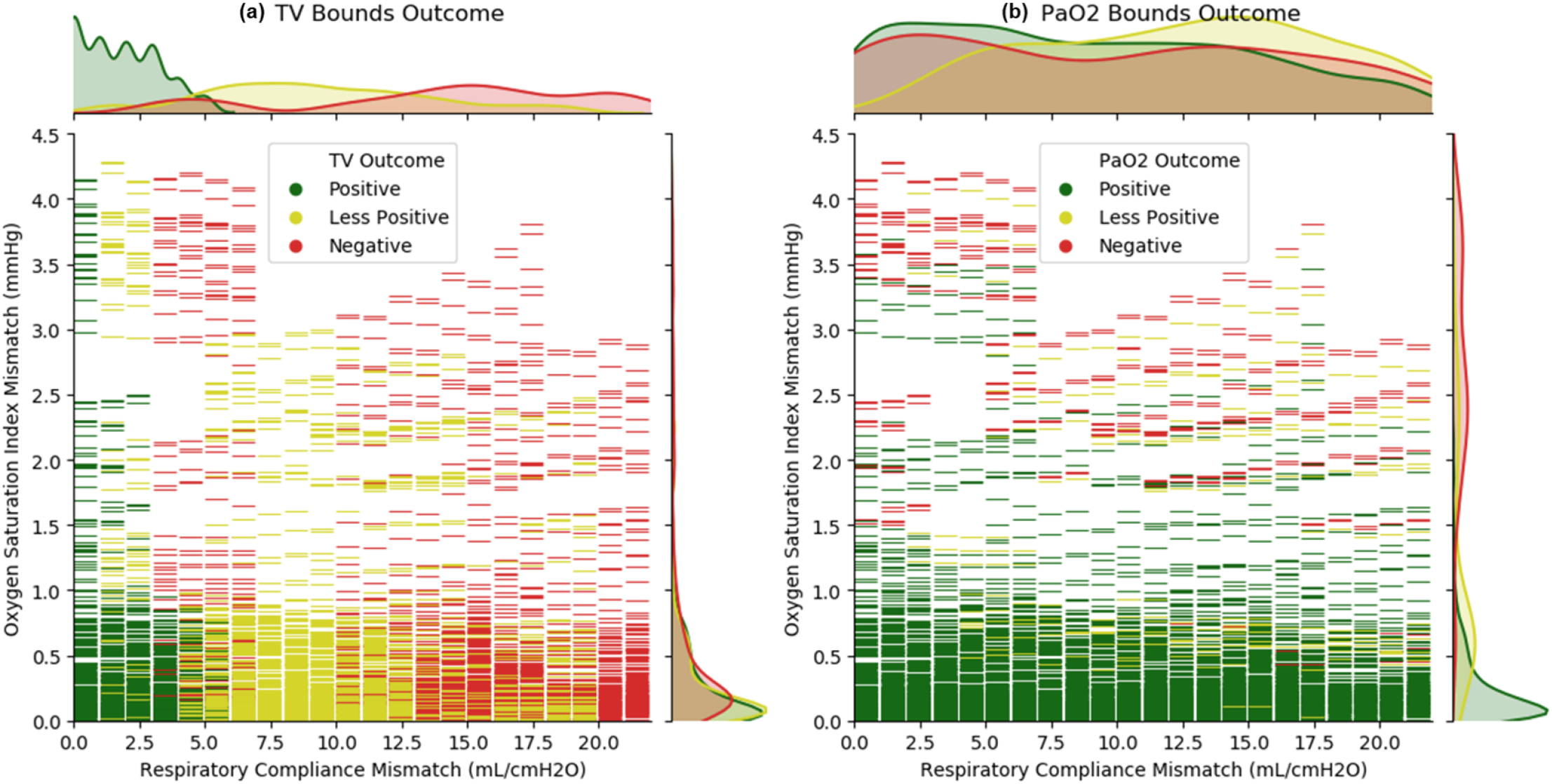
Comparison of multi-patient ventilation simulated outcomes due to TV (plot a) and PaO_2_ (plot b) outcome bounds. Each graphical dash is a full simulation. Included are univariate histogram plots for each axis using kernel density estimation to represent the distribution of all three outcomes described in two dimensions. The compliance (abscissa) has discrete values due to the chosen patient model parameter setting methodology and fluid mechanics. The OSI (ordinate) is dependent on all external settings, along with the complex interactions of internal mechanistic models. Note that while the OSI has units of mmHg (because it is a ratio of pressure divided by saturation), the interpretation is like the unitless OI value.

While respiratory compliance is a direct indicator of outcome based on lung recruitment, Fig 5 shows that OSI is an important and effective measure of overall diffusion impairment.

**Fig 5.**
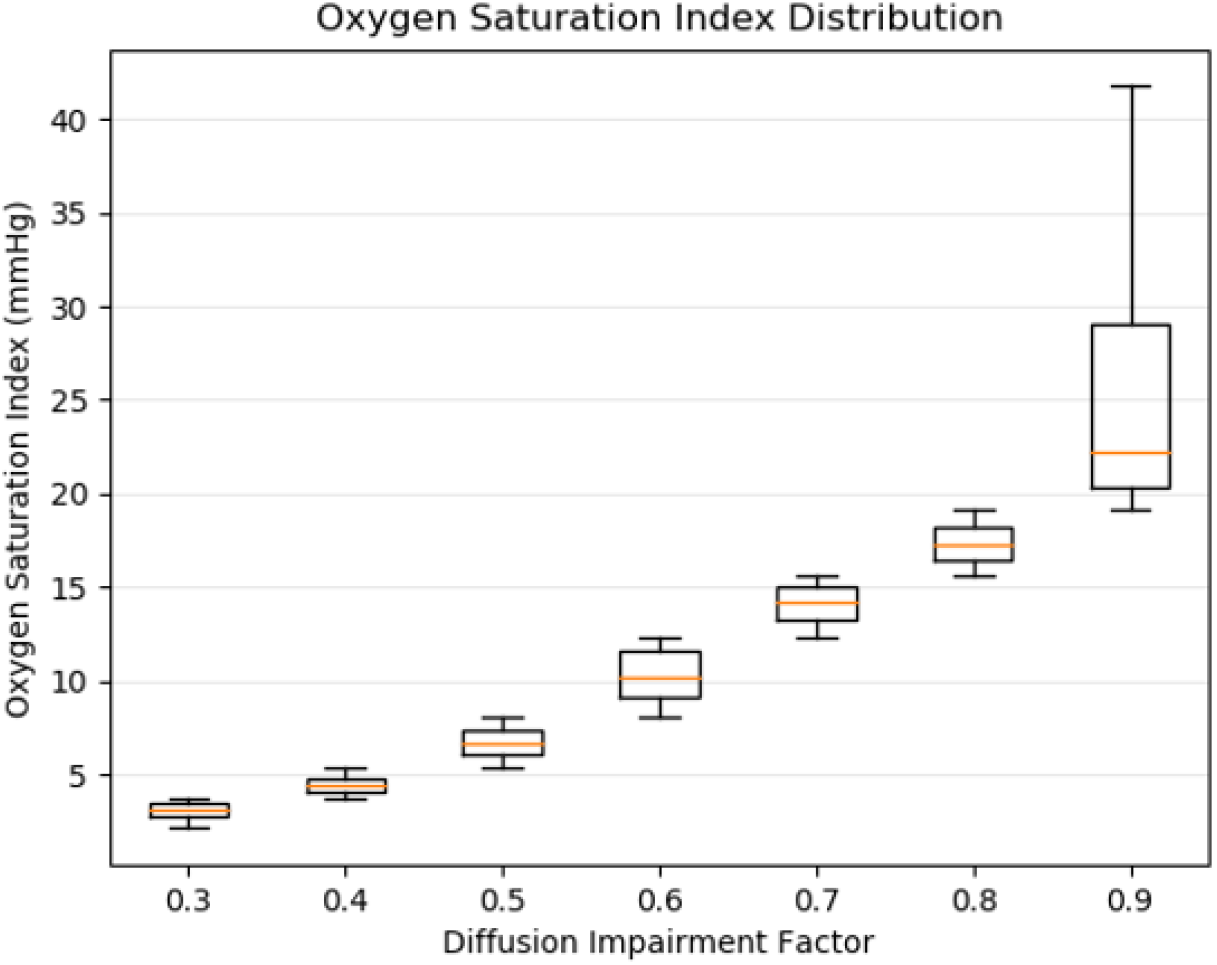
Distribution of all simulated patient’s OSI grouped by the seven DIF settings used from mild to severe. The OSI increases with DIF and is, therefore, useful for non-invasive clinical assessment of hypoxemic respiratory failure. The OSI increases with diffusion impairment because the SpO_2_, which plateaus at 100, is a proportionately larger fraction of PaO_2_ as diffusion impairment and shunt make PaO_2_ less than expected for a given FiO_2_.

The combined outcomes as a function of the patient parameter mismatches are used to generate a complete decision matrix in Fig 6. The simulation results show that patients with similar respiratory compliances and comparable OSI are most likely to have satisfactory outcomes when paired to a single ventilator.

**Fig 6.**
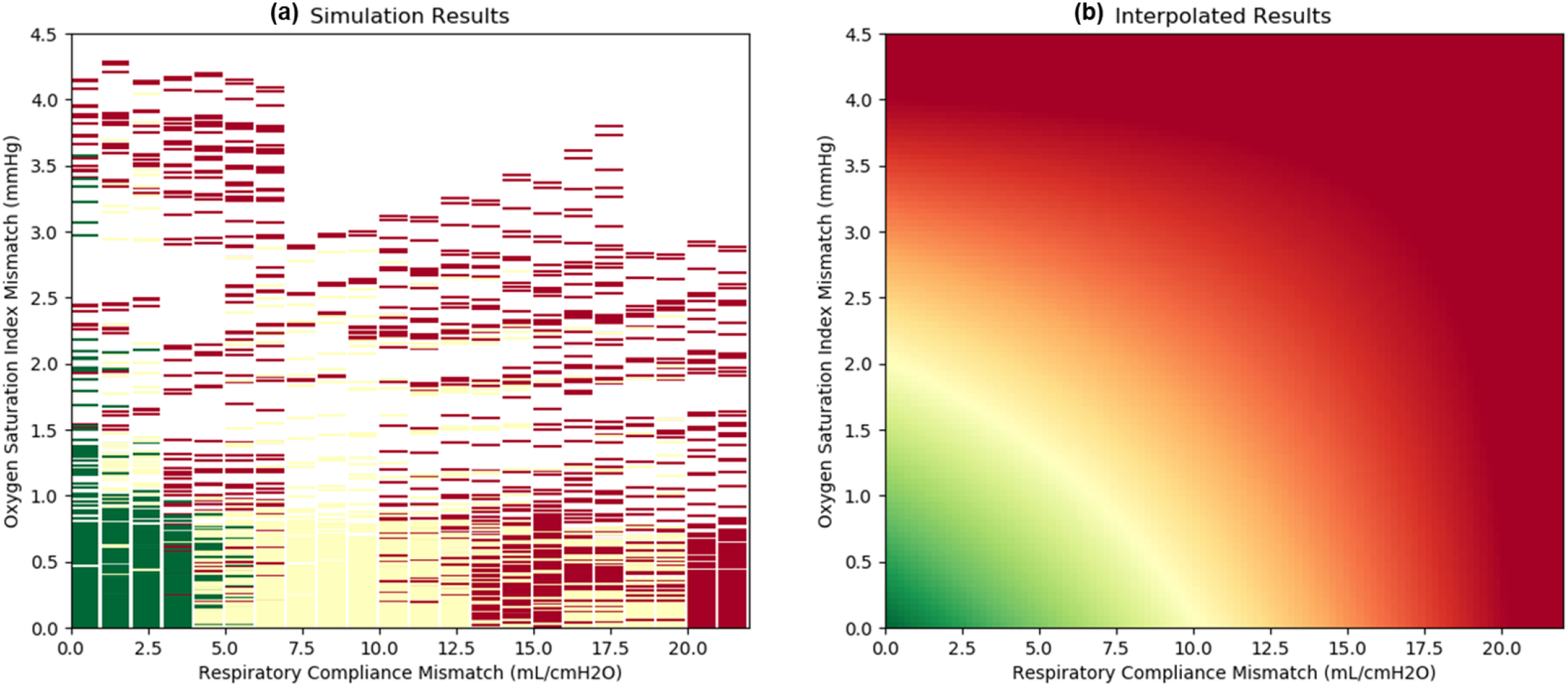
The simulations from Fig 4 were holistically taken into account to get a complete decision matrix. Outcomes were assigned a normalized value of green=1, yellow = 0.5, and red = 0 to encode a z-axis as colors or color gradient. The resulting three-dimensional scatter plot (plot a) was used to produce an interpolated surface using the first-order bivariate B-spline method (plot b).

An interactive version of the model is available via a Jupyter notebook in the Pulse repository (https://gitlab.kitware.com/physiology/jupyter).

## Discussion

The results demonstrate that lung compliance and OSI are useful clinical guidance parameters for multi-patient ventilation, as they are non-invasively measurable and highly correlated to outcomes. OSI has also been shown to be a significant indicator of clinical outcomes for ARDS [39]. Our simulations show that a difference up to approximately 12 mL/cmH_2_O in lung compliance and an OSI difference of less than 2 mmHg anticipate satisfactory patient outcomes. (While the OSI has nominal units of mmHg as an artifact of its calculation, it is really to be interpreted as a ratio of oxygen partial pressure to hemoglobin saturation, which is unitless. Conveniently, oxyhemoglobin saturation of 100% typically corresponds to about 100 mmHg oxygen partial pressure for normal lungs.) The range of simulated parameter variation is more liberal than the conservative protocol of Beitler et al. [18] With a simple circuit, there is less flexibility than may be achieved with the more complex implementation proposed by Han et al. [23] This simulation may inform clinical guidelines when pairing patients on a single ventilator. An obvious limitation of this conclusion is that this is a pure simulation study, and we do not have correlated patient data. Planned future work includes comparing our computational simulations to physical experiments using the simple lung simulators with discrete selectable settings for resistance, compliance, and effects of negative inspiratory pressure patient-initiated breath triggers [IngMar Medical, QuickLung® Adult Precision Test Lung with QuickTrigger, part number 15 00 100, www.ingmarmed.com]. Parameters of this simulation model were chosen to match nominal QuickLung® parameters to facilitate future bench testing. Clinical validationmay never be possible, would likely be observational at best, and is very unlikely to undergo ethical randomized clinical trial. This is a value of simulation, that possibilities can be explored that are beyond the realm of clinical investigation, yet of clinical value.

Implementation of split breathing circuits without compensating resistances was the specific model addressed in the joint statement by professional societies recommending against this practice [13]. Various means have been employed empirically to limit flow to the most compliant lungs [19-23]. Modeling flow restriction adds model parameters, greatly increasing the number of simulations needed. For those locales with the most rudimentary resources, flow restriction devices may not be possible. Future work will also investigate sharing ventilators by more than two patients, and the use of flow restrictors in circuit branches to facilitate separate control of tidal volume delivered to individual patients during multi-patient ventilation.

If there is an adequate supply of ventilators available, individual ventilators are preferable and optimal [5,7,12]. Numerous efforts to produce quantities of individual ventilators have been kindled by an anticipated surge of patients needing ventilator support. There has been a surge of creative projects, yet there are gaps to be bridged before mass production could be a reality [40]. The FDA has issued Emergency Use Authorizations (EUA) for some innovative ventilators not made by usual medical equipment vendors [41]. Whether a surge of supply will be needed or available remains to be seen.

However, even with numerous creative projects creating basic ventilators from readily available components, there may not be enough locally available in a surge emergency. The primary default is manual ventilation, usually with bag-valve-mask (BVM) devices. Prolonged manual ventilation is difficult to maintain. [42] Split breathing circuits can be quickly assembled from available breathing circuit components for a transition to mechanical ventilation. Ventilation equipment not usually employed for critical care may also be pressed into service in a crisis, e.g., CPAP and BiPAP as ventilators, anesthesia machines, and high-flow non-invasive ventilation devices.

Multi-patient ventilation may be the only available option for poorly resourced regions. Manufacturing of single patient ventilators, however primitive, is an unlikely prospect for countries with limited resources. A personal communication from a Sudanese engineer interested in producing ventilators in-country stated that the entire country has only 80 ventilators and very limited materiel or manufacturing resources. Whatever can be done with resources already present in the country is their only available option (Conversation with MS, 28 Mar 2020, SMP). Similar limitations also apply for very populous countries, such as India, though they may be able to rally more resources for high throughput manufacturing. For these critical scenarios, we intend that these simulations could inform clinical guidelines for patient pairing on a single ventilator.

Under extreme circumstances, and in resource-challenged regions limited by the availability of an adequate number of ventilators, multi-patient ventilation is a potentially viable option and can significantly increase the capacity to care for critically ill patients in surge scenarios. Our computational simulation study provides ranges of basic physiological parameters that could be used in patient selection for future studies evaluating multi-patient ventilation. As a trade-off under urgent conditions, this simulation suggests that less stringent matching criteria may be tolerable without imposing a very deliberate and conservative protocol [18]. However, implementation would be less tolerant of patient variations and faults than the more complicated arrangement of partially independent patient breathing circuits [23]. Our simulation could inform clinicians acutely needing to support two patients with a single ventilator. Further physical validation of these simulation results is needed.

Inevitably, ethical issues arise. Some will question whether it is ethical to undertake a desperate ventilator sharing tactic without a well defined exit strategy. This is not unlike many clinical interventions that secure time for informed triage and more deliberate consideration of longer term options. However, sharing a ventilator has implications for more than a single patient. Response to an epidemic challenge may foster innovation with broad implications, for instance extensive use of manual ventilation of polio patients in 1952 that kindled vigorous development of mechanical ventilators [43] and the discipline of critical care. We regard uncompensated ventilator sharing as a desperate “entrance strategy”. The underlying question is whether it is unethical to maintain ventilation with imperfect apparatus as an alternative to not providing ventilation at all, despite not having a long-term exit strategy. This conundrum is worthy of careful and nuanced discussion, yet beyond the scope of this technical study. As in Las Vegas, this approach might be mitigation while single-patient ventilators are in transit from a remote location. It could be an intermediate or longer term intervention. These are unexplored considerations forced upon us by advent of a global pandemic. The variety of possible surge scenarios defies adequate planning for every contingency. [42,44-46] Perfect can be the enemy of good. The best possible action, though not optimal, is not necessarily futile. [1,2,47]

The simulation software, data, and a UI for exploring the simulations is available open-source at the Kitware GitLab site (https://gitlab.kitware.com/physiology). For more details, see the supplement.

## Conclusion

### Evidence before this study

If numbers of patients requiring mechanical ventilation exceed the number of available ventilators in a surge, shared branched ventilator circuits have been proposed for sharing one ventilator by multiple patients. Under pressure of imminent pandemic surge, only small and rudimentary clinical studies were available. Testing over expected ranges of lung-chest wall compliance were not found. Increasing clinical experiences of mechanical ventilation parameters employed for COVID-19 patients are being reported.

### Added value of this study

The number of possible combinations of ventilation and physiological parameters is very large. Time and resource constraints do not permit conventional research. Computational simulation provides rapid sensitivity evaluation of several factors over a wide range of hypothetical ventilation conditions. Envelopes of evaluated parameters may provide reasonably estimated safety boundaries for clinicians compelled in an emergency surge to employ a poorly characterized practice. A previously well-vetted computational model for ventilation of a single patient by a dedicated ventilator has been modified to model the sharing of a single ventilator by two or more patients. Only pairings of two equally sized 70 kg patients are modeled in this report. These simulations provide estimates of effects on ventilation and blood oxygenation by clinically measurable values using conceivable mismatched patient lung compliance and oxygenation (diffusion and shunt).

### Implications of all the available evidence

These estimates are for pressure mode ventilation using a single ventilator shared by branched breathing apparatus for two patients. Individual patient flow restriction to compensate for compliance mismatch is not considered in this simulation. Reasonable though arbitrary bounds of acceptable parameters may guide clinicians when evaluating candidate pairings of patients with different physiological characteristics. Further laboratory testing and clinical experience will be needed to determine the validity or utility of these assessments. Different simulations will be needed for flow-compensated branches, more than two patients, and unmatched body habitus.

## Data Availability

All data is available as open source (Apache License, Version 2.0).

https://gitlab.kitware.com/physiology/engine/-/blob/3.x/src/cpp/study/multiplex_ventilation/ReadMe.md

https://gitlab.kitware.com/physiology/engine/-/tags/STUDY_MULTIPLEX_VENTILATION_1_0_0

https://gitlab.kitware.com/physiology/jupyter

## Supporting information

### Multi-patient ventilation design

The lumped-parameter models in the Pulse Physiology Engine are shown in S1 Fig for the respiratory system and the ventilator. Two patients respiratory circuits are connected to a single ventilator for transient circuit analysis. The air pressure, flow, and volume and substance amounts are calculated for each time step. This physics-based approach provides an flexible methodology for changing the resistors and/or capacitors in the lumped-parameter models to represent disease states, such as ARDS.

**S1 Fig.**
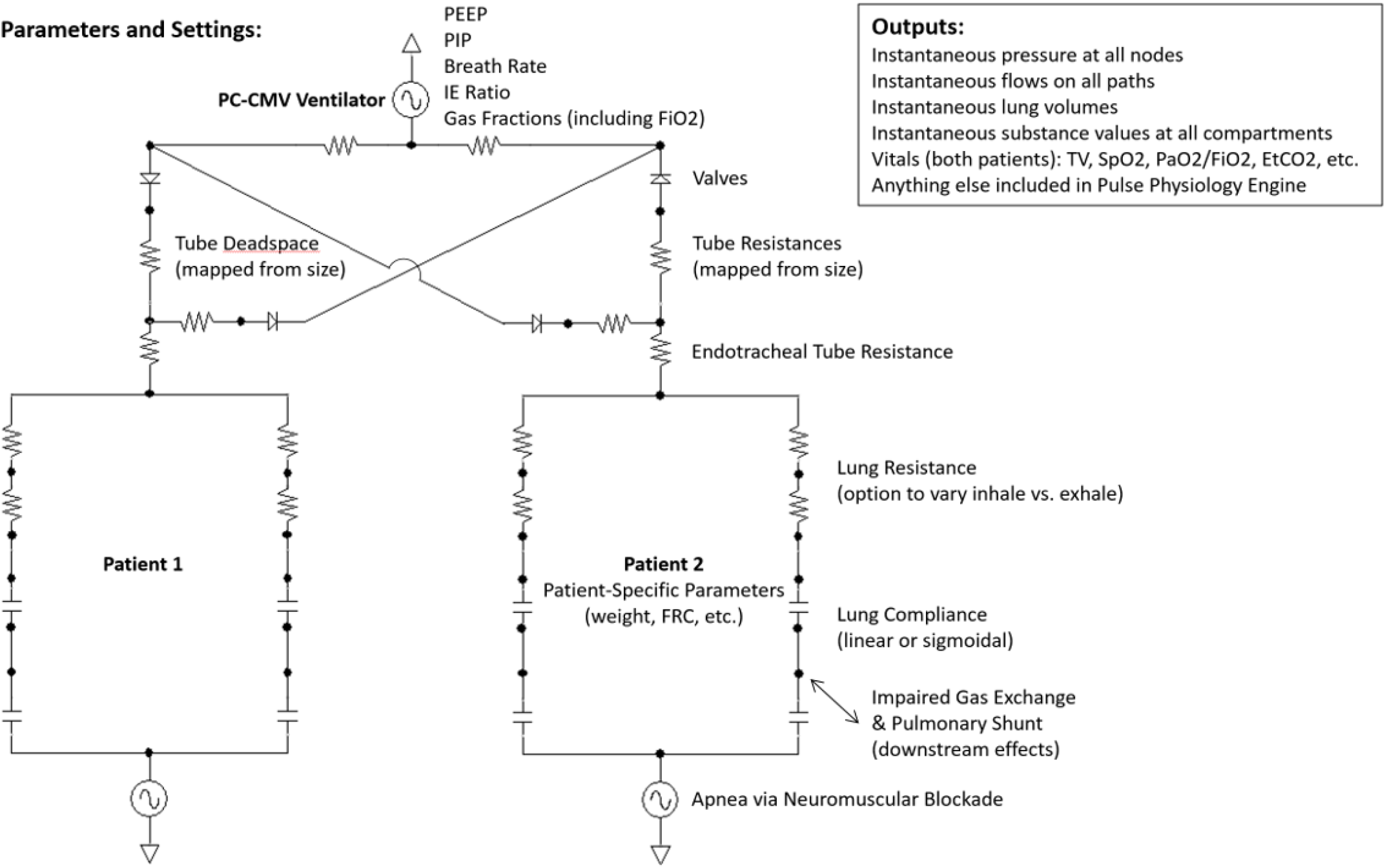
The combined closed-loop lump-parameter fluid circuit for simulating multi-patient ventilation. A new entire system state is calculated every two ms. The Pulse dynamic circuit solver and transporter is leveraged to ensure sound physics-based results with conservation of energy and mass. Not explicitly shown are interactions with all other Pulse physiological systems, most notably, the alveolar-capillary partial pressure gradient diffusion gas exchange with the cardiovascular system.

### Diffusion impairment factor design

To implement the COVID-19 disease state, the Diffusion Impairment Factor (DIF) was calculated. The factor was mapped to the cardiovascular circuit in Pulse at the intersection of the cardiovascular and respiratory circuits, as shown in S2 Fig.

**S2 Fig.**
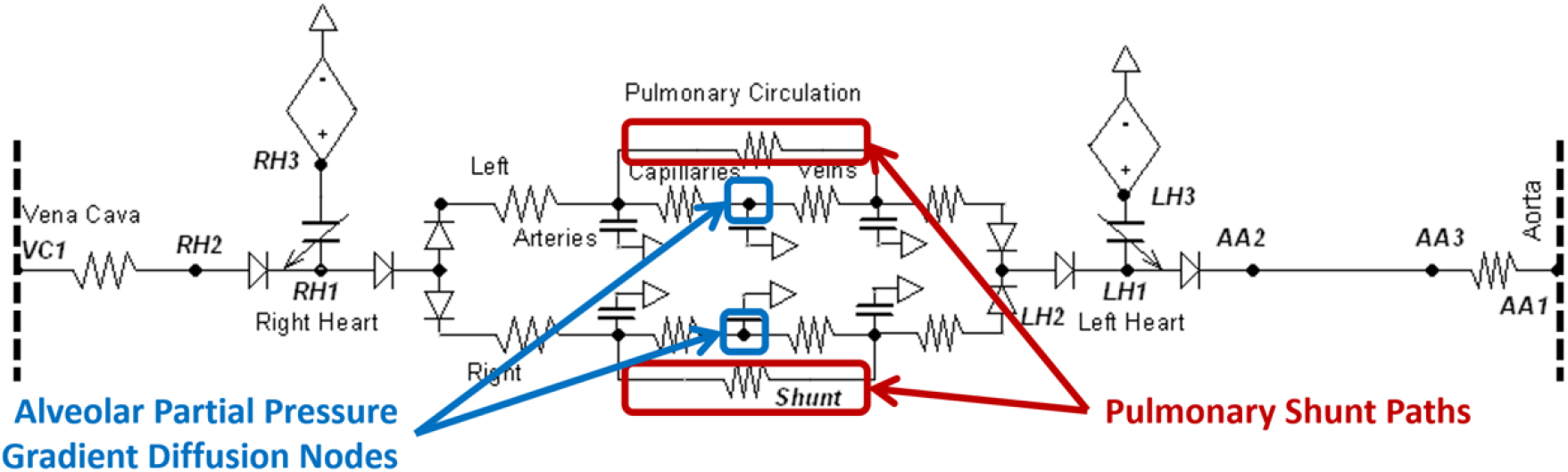
The Diffusion Impairment Factor (DIF) implementation on the pulmonary section of the cardiovascular circuit in the Pulse Physiology Engine. The DIF severity is mapped to cardiovascular circuit modifiers to hinder alveolar gas exchange and increase pulmonary shunting.

The pulmonary shunt resistance and alveolar surface area are both reduced by a factor (y) based on severity (x [0 to 1]) following the exponential decay equation

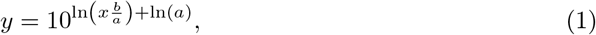

Where, a is 1.0 and b is 0.1 for the alveolar surface area and a is 1.0 and b is 0.01 for the pulmonary shunt resistance change.

### Patient exclusion criteria

As noted in the Methods section, the patients were initially stabilized on a single ventilator. By varying the DIF and modifying the compliance, the patients were created with various disease states. Some of these patients were unable to meet our scoring criteria for multi-patient ventilation on an individual ventilator. Therefore, they were excluded from further analysis, so prevent noise in the data. S1 Table shows the percentage of patients able to meet the oxygen saturation scoring criteria of at least 89%.

**S1 Table.** Patient Exclusion Results. Patients with higher DIF severities require a higher ventilator FiO_2_ setting to achieve positive (green) outcomes based on SpO_2_. The total fraction of patients that are included for multi-patient simulation analysis are listed in the third column. The minimum FiO_2_ setting is 0.21 (natural air).

| DIF | Mean Minimum $\text{FiO}_2$ for $\text{SpO}_2 > 89\%$ | Fraction of Patients Achieving $\text{SpO}_2 > 89\%$ |
| --- | --- | --- |
| 0.3 | 0.21 | 100% |
| 0.4 | 0.21 | 100% |
| 0.5 | 0.23 | 100% |
| 0.6 | 0.27 | 100% |
| 0.7 | 0.46 | 100% |
| 0.8 | 0.95 | 29% |
| 0.9 | N/A | 0% |

### Simulation pseudocode

For this study, we chose the following ranges (Minimum, Maximum, Step):

- PEEP (cmH_2_O): Minimum:10, Maximum:20, Step Size:5
- Compliance (mL/cmH_2_O): Minimum:10, Maximum:50, Step Size:10
- DIF (%): Minimum:30, Maximum:90, Step Size:10

This generates a total of 12,642 unique simulations to execute. The execution loop is shown in Algorithm 1.

#### Algorithm 1 The Pulse multi-patient execution pseudocode for this investigation

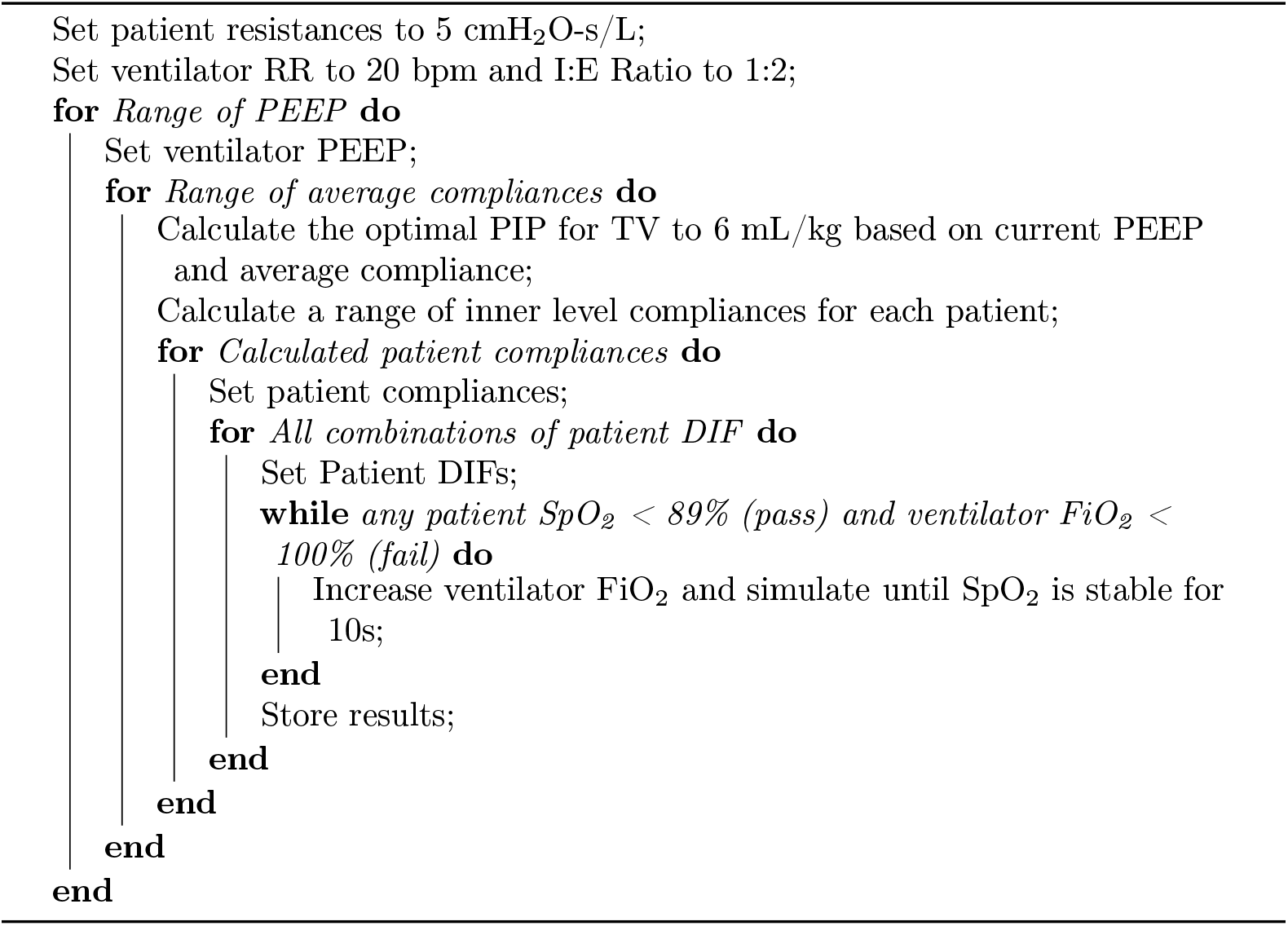

Oxygen saturation stability is determined by storing the lowest SpO_2_ of all engines as a baseline value at a specified time. For each subsequent time step, we sample the lowest SpO_2_ calculated and determine the percent difference with the baseline value. The SpO_2_ is considered stable when the percent different is less than 0.25% for 10s. Once a stable SpO_2_ value is achieved, the FiO_2_ setting on the ventilator is adjusted accordingly:

- If the SpO_2_ is trending down and < 85%, FiO_2_ is increased by 10%
- If the SpO_2_ is trending down and ≥ 85%, FiO_2_ is increased by 2.5%
- If the SpO_2_ is trending up and < 88%, FiO_2_ is increased by 5%
- If the SpO_2_ is trending up and > 88%, FiO_2_ is increased by 1%

### Pulse Physiology Engine architecture

Architecturally, a single instance of the multiplex ventilation engine accepts a list of simulations to run. Each simulation in the list contains the compliance, resistance, and impairment for each patient, and the ventilator settings for the simulation. As each engine simulates, it creates a log of the simulation and a CSV (Comma Separated Values) file containing the simulation results for each timestep. When both patients within the engine reach final homeostasis, the final values of interest will be appended to the simulation list for the entire experiment. When all engines have completed, the simulation list is output as a JSON (JavaScript Object Notation) file.

### Pulse Physiology Engine data model

Pulse implements its data model (model definitions) via Google Protocol Buffers (GPB). This provides the ability to support instantiation and control of Pulse engines from various programming languages and network protocols. GPB also provides the ability to read and write data in the popular JSON format for readability and import into popular data analysis libraries. The GBP definitions used by the Multiplex Ventilation Engine can be found here, and below is an example of the data properties computed for a single patient.

Patient State JSON:

{

“ID”: 0,

“Compliance_mL_Per_cmH2O”: 10,

“Resistance_cmH_2_O_s_Per_L”: 5,

“ImpairmentFraction”: 0.40000000596046448,

“RespirationRate_Per_min”: 20,

“IERatio”: 0.5,

“PEEP_cmH_2_O”: 10,

“PIP_cmH_2_O”: 55,

“FiO_2_”: 0.21,

“AirwayFlow_L_Per_min”: −384.06825814934518,

“AirwayPressure_cmH_2_ O”: 1043.4194836658687,

“AlveolarArterialGradient_mmHg”: 45.619370140146856,

“ArterialCarbonDioxidePartialPressure_mmHg”: 39.819789561334652,

“ArterialOxygenPartialPressure_mmHg”: 75.85119486614299,

“CarricoIndex_mmHg”: 367.5726868121024,

“EndTidalCarbonDioxidePressure_mmHg”: 20.822935895891064,

“IdealBodyWeight_kg”: 75.3,

“MeanAirwayPressure_cmH_2_O”: 25,

“OxygenationIndex”: 11.003491209618154,

“OxygenSaturation”: 0.9603982517625641,

“OxygenSaturationIndex_mmHg”: 6.3974838958631794,

“SFRatio”: 4.6503130439307032,

“ShuntFraction”: 0.087940097305858056,

“TidalVolume_mL”: 449.99997579831688,

“TotalLungVolume_mL”: 2680.9772230837052,

“AchievedStabilization”: true,

“OxygenSaturationStabilizationTrend”: −0.11851467873102428

}

### Resource requirements

Pulse has a small memory footprint (< 10MB of RAM) and simulates at 10x real-time while using 100% of one average CPU core. Since the multiplex ventilation engine contains two patients, but with a shared respiratory circuit it runs around 6x real-time using 100% of one CPU core. Pulse is not a good candidate for multi-threaded execution of the physiology analysis as the individual system models must be executed in series. A runner was created to run each simulation in the simulation list on its own CPU core via threads. For our study, we employed an Alienware Area-51 R6 with dual processors (16-Cores each, overclocked up to 3.6GHz) and 64GB of RAM.

### Graphical user interface

As part of this effort, we created bindings to instantiate and control the multiplex engine using the Python programming language. We were able to create and deploy a Jupyter Notebook that provides a simple interface for end users to customize their own patients (via compliance, resistance and impairment factors) and control the ventilation while viewing simulation results in real-time. The Jupyter Notebook GUI can be found in our gitlab repository (https://gitlab.kitware.com/physiology/jupyter) and is shown in S3 Fig.

**S3 Fig.**
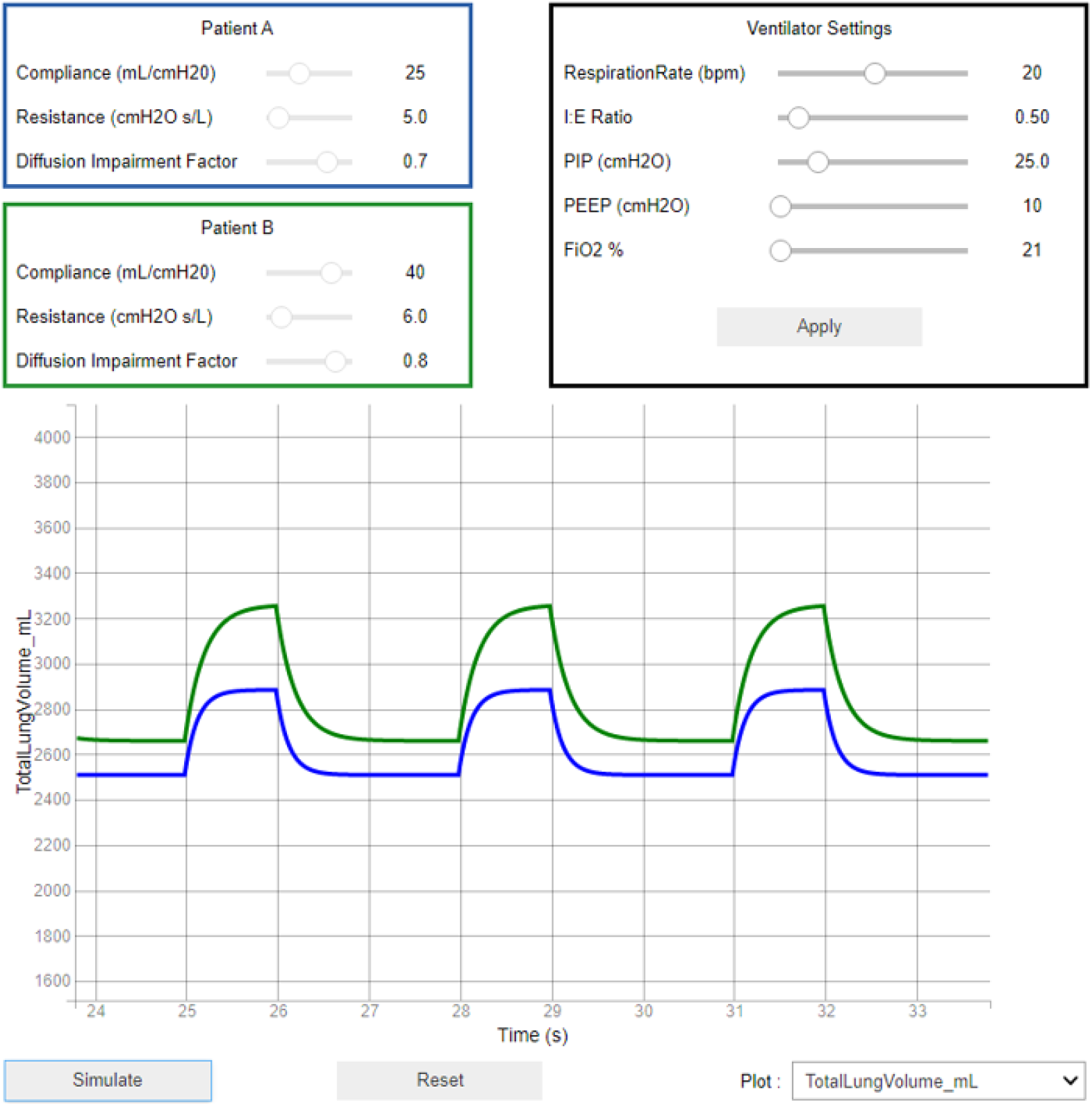
Jupyter Notebook User Interface. The user interface allows for patient customization of the disease state and the ventilator settings for multi-patient ventilation. Results of the simulation are shown in the lower portion of the simulation.

### Source code and associated data

The Pulse Physiology Engine, the multiplex ventilation engine for Pulse, and the GUI are open source under the Apache 2.0 software license. All code can be obtained from our gitlab repository (https://gitlab.kitware.com/physiology/engine/-Ztags/STUDY_MULTIPLEX_VENTILATION_1_0_0), and can be used to generate all of the final study data. To build the engine, follow the build instructions provided on the repository ReadMe: https://gitlab.kitware.com/physiology/engine/-/blob/3.x/ReadMe.md All data used in this study, as well as run instructions for the multiplex ventilation engine can be found in the multiplex ventilation engine ReadMe: https://gitlab.kitware.com/physiology/engine/-/blob/3.x/src/cpp/study/multiplex_ventilation/ReadMe.md.

